# Predicting COVID-19 (Coronavirus Disease) Outbreak Dynamics Using SIR-based Models: Comparative Analysis of SIRD and Weibull-SIRD

**DOI:** 10.1101/2020.11.29.20240564

**Authors:** Ahmad Sedaghat, Amir Mosavi

## Abstract

The SIR type models are built by a set of ordinary differential equations (ODE), which are strongly initial value dependant. To fit multiple biological data with SIR type equations requires fitting coefficients of these equations by an initial guess and applying optimization methods. These coefficients are also extremely initial value-dependent. In the vast publication of these types, we hardly see, among simple to highly complicated SIR type methods, that these methods presented more than a maximum of two biological data sets. We propose a novel method that integrates an analytical solution of the infectious population using Weibull distribution function into any SIR type models. The Weibull-SIRD method has easily fitted 4 set of COVID-19 biological data simultaneously. It is demonstrated that the Weibull-SIRD method predictions for susceptible, infected, recovered, and deceased populations from COVID-19 in Kuwait and UAE are superior compared with SIRD original ODE model. The proposed method here opens doors for new deeper studying of biological dynamic systems with realistic biological data trends than providing some complicated, cumbersome mathematical methods with little insight into biological data’s real physics.

## I. Introduction

Since the outbreak of the novel coronavirus (COVID-19) in Wuhan, China in December 2019, the world has experienced the worst pandemic in history; not in terms of fatality but in the way we have used to live and commute around world. Many researchers and scientists from different discipline have made efforts to better understand and manage the COVID-19 pandemic. On 15 June 2020, COVID-19 infectious was reported 8,066,465 cases with 437,295 deaths and 4,174,782 recovered in 215 countries worldwide [1]. Accurate prediction of COVID-19 dynamics is highly crucial for the governments and health organizations to better management of the situation. Exact solutions of SIR model were reported in a number of publications; although, these solutions hardly fit with actual data from a pandemic. Bohner et al. [2] provided a nice article on Bailey’s [3] SIR model’s exact solution. In such methods, the recovered (R) population is ignored when solving susceptible (S) and infectious (I) equations; hence, we couldn’t fit the COVID-data with the explicit formulations provided. Harko et al. [4] have reported exact solutions to SIR model considering birth and death rates; yet no actual epidemic data were tested with these solutions. Shabbir et al. [5] and Maliki [6] have both reported exact solutions to SIS and SIR original ODE equations reported by Kermack and McKendrick [7]. These special models included (S) and (I) equations only.

In this paper, we consider SIRD model, which considers susceptible (S), infected (I), recovered (R), and deceased (D) ordinary differential (ODE) equations. Providing a formulation for infectious population using the Weibull function, we can easily find all explicit solutions to the rest of the equations without the need to solve ODE. The present formulation provides fast and robust solutions to SIRD equations and can be used to optimize the highest prediction of an epidemic/pandemic. The goodness of fit to the Weibull-SIRD model for COVID-19 data in Kuwait and UAE is examined using the determination coefficient (R2). Results of COVID-19 dynamics are discussed for Kuwait and UAE, and the prediction capability of the new model is presented, and conclusions are drawn.

## II. Materials and methdos

A fast and robust mathematical model solution to any new pandemic outbreak such as COVID-19 is required to reduce disastrous consequences and equip with appropriate measures. COVID-19 indicated the gap on human fragility in tackling biological crisis. Here, we first provide the formulation of SIRD model base on susceptible, infected, recovered, and deceased population [8, 9].

### A. SIRD Model

In epidemiological studies, it is assumed that total population size is not changing and remain constant during epidemy. Human factors such as sex, age, social behaviour, and location are not considered in SIR model which is not completely correct. In COVID-19 pandemic, it is observed that transmission rate is strongly dependent on social behaviour and habits of individuals. The SIRD model consist of 4-set of ordinary differential equations (ODE) for susceptible population (S), infected cases (I), and recovered cases (R), and deceased cases (D) as follows [10, 11]:

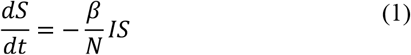

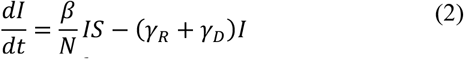

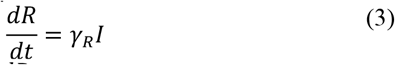

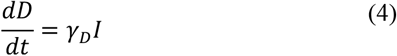

Initial total population (*N*) is assumed to be constant during pandemic as follows:

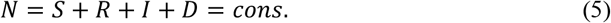

In the above equations, the transmission rate (*Β*) shows growth of an infected epidemic. The recovery rate (*γ*_*R*_) shows growth of recovered cases and the death rate (*γ*_*D*_) indicates deceased cases during a pandemic. The removing rate (*γ* = *γ*_*R*_ + *γ*_*D*_) expresses recovered and deceased populations removed from susceptible population. Another important factor is called the reproduction number (*R*_0_= *β*/*γ*) dictating outbreak of an endemic. If *R*_0_> 1 then it is expected an infected person comes in contact of *R*_0_susceptible people before he/she is removed.

As observed in above equations, if the solution to equation (2) on infected population is known, then we can easily solve recovered and deceased equations (3) and (4). Below, an analytical solution based on Weibull distribution function is introduced.

### B. Weibull-SIRD Model

The most commonly used Weibull distribution function of two parameter family (*k, c*) may be reformulated for infected population of a pandemic as follows [12]:

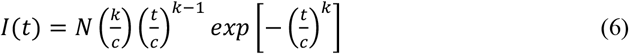

In equation (6), *I*(*t*) is the Weibull function of daily infected population, *N* is the total susceptible population, *k* is a unitless shape factor and *c* is a constant time on unit of (day), usually referred as scale factor. The values of *N, k*, and *c* are model parameters and are obtained by fitting Weibull function to infectious population (I) from a pandemic data.

Exact solutions to recovered (R) and deceased (D) is simply obtained by plugging equation (6) into equations (3) and (4) to obtain cumulative solution as follows [12].

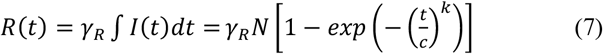

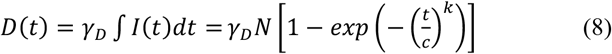

Substituting equation (6) into equation (1), one may obtain a solution for susceptible population as follows.

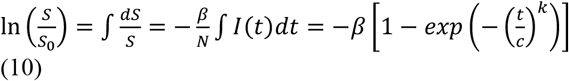

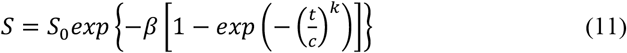

By changing the total susceptible population (N) in SIRD model, the recovered and deceased will be massively changed. However, in Weibull-SIRD model, we should stick to a fixed number for N to best fit infected cases with Weibull function in equation (6). To fine-tune the expected recovered and deceased cases, we propose to use variable recovery rate (*γ*_*R*_) and deceased rate (*γ*_*D*_) as follows.

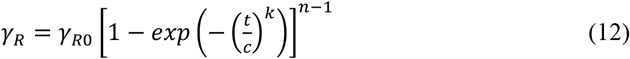

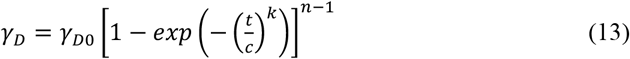

The power factor (n) is any number and can be adequately fine-tuned to best fit the recovered and deceased populations data.

The above set of analytical equations (6), (7), (8), and (11) provide a simple and robust solution to an endemic/pandemic data. We have applied optimization to get best value of coefficients in SIRD and Weibull-SIRD models. MATLAB algorithm (lsqcurvefit) was applied to find best coefficients in SIRD model equations (1) to (5). MATLAB algorithm (fminsearch) was used to find best fit coefficients in Weibull-SIRD model equations. The goodness of fitted COVID-19 data are examined using the coefficient of determination (R^2^) [13].

## III. Results

Results of Weibull-SIRD analytical model are provided and discussed for COVID-19 in Kuwait and UAE below.

### A. COVID-19 Prediction in Kuwait

Results of Weibull-SIRD analytical model are provided and discussed for COVID-19 in Kuwait and UAE below.

The results of fitting equations (6) - (13) with COVID-19 data in Kuwait are given in Table 1.

Figure 2 shows that 4-set of cumulative COVID-19 data including susceptible, infected, recovered, deceased are simultaneously fitted with Weibull-SIRD curves. Goodness of fit (R^2^) values are shown in Table 1. As shown in Fig. 2, on Sunday 31 May 2020 Kuwait had passed the peak of COVID-19 infectious with 14,814 infected cases. Both peak dates and values reported 10 June 2020 and 18, 431 using direct solution of SIRD ODE equations [15,16]. COVID-19 data for Kuwait on 31 May 2020 suggest 15,445 infected cases; while on 10 June 2020, there was 10,260 infected cases. Weibull-SIR model predicts 10,581 infected cases on 10 June 2020. It is obvious from the results that the Weibull-SIR model provided closer and robust solution to COVID-19 pandemic in Kuwait.

**Table 1:** Optimized Weibull-SIRD parameters for COVID-19 in Kuwait (18 June 2020).

| Total population (N) | shape factor (k) | Scale factor (c) | Growth rate ($\beta$) | Recovery rate ($\gamma_{R0}$) | Death rate ($\gamma_{D0}$) | Removal rate ($\gamma_0$) | Reproduction number ($R_0$) | $R^2(S)$ | $R^2(I)$ | $R^2(R)$ | $R^2(D)$ |
| --- | --- | --- | --- | --- | --- | --- | --- | --- | --- | --- | --- |
| 555,509 | 7.1 | 98.97 | 0.0786 | 0.0686 | 0.00083 | 0.02719 | 1.13 | 0.9855 | 0.975 | 0.9495 | 0.7193 |

**Figure 1:**
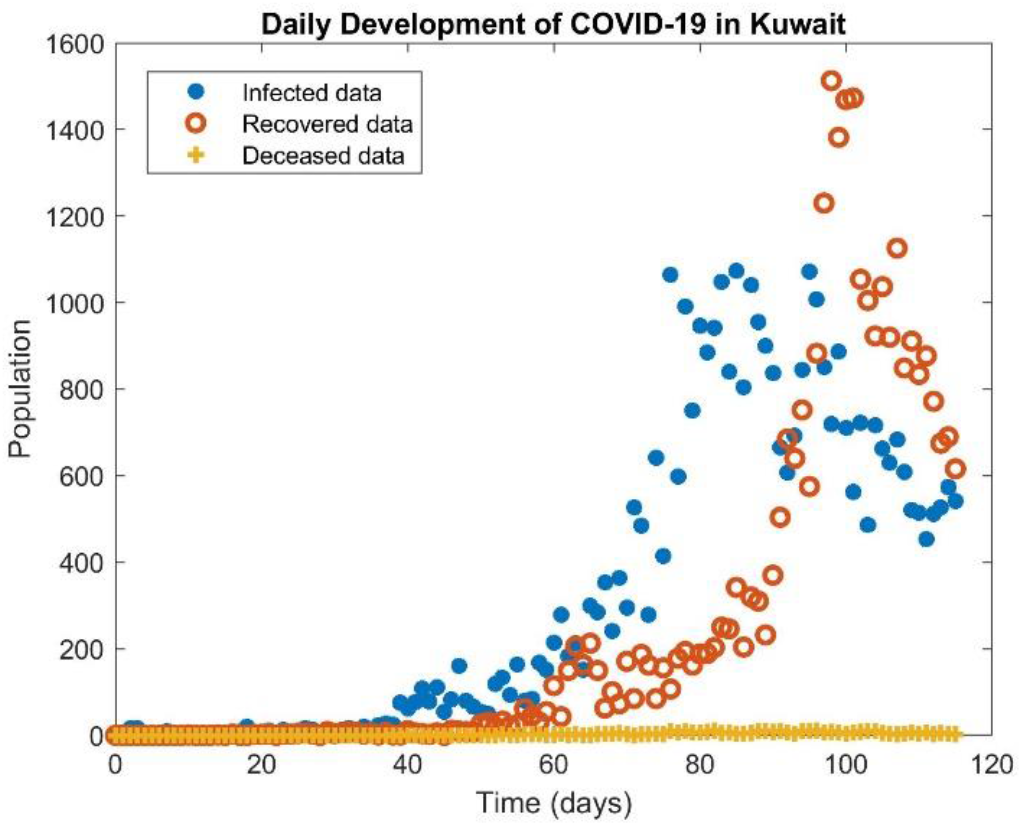
Daily exposure of COVID-19 pandemic in Kuwait

**Figure 2:**
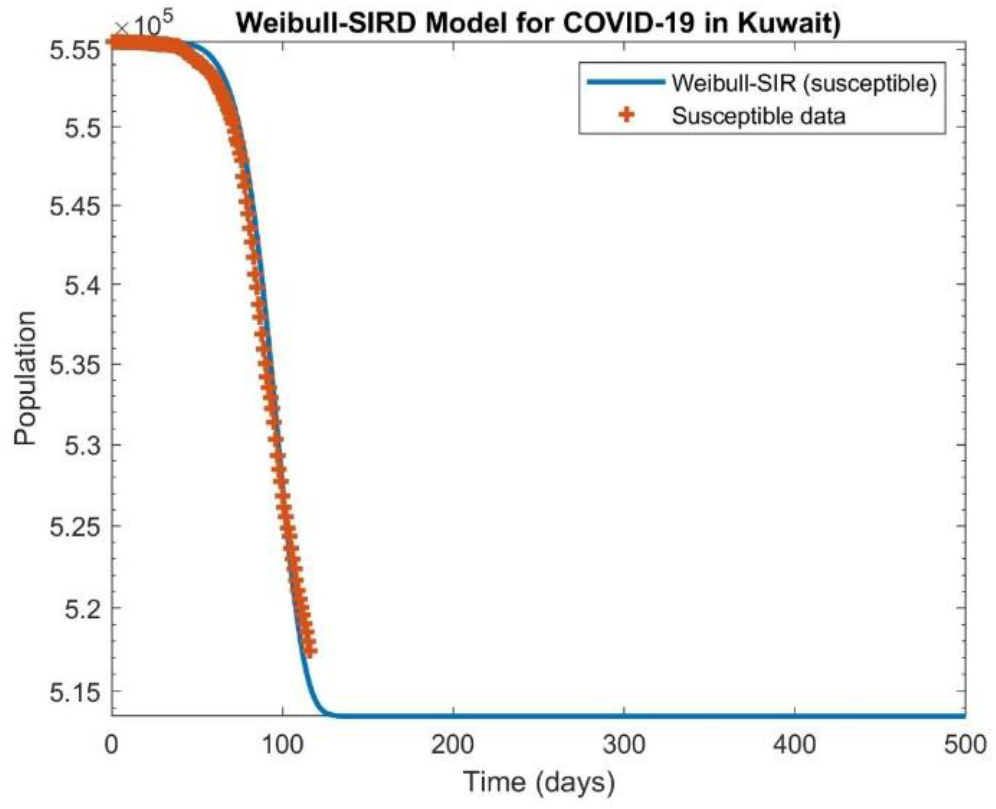
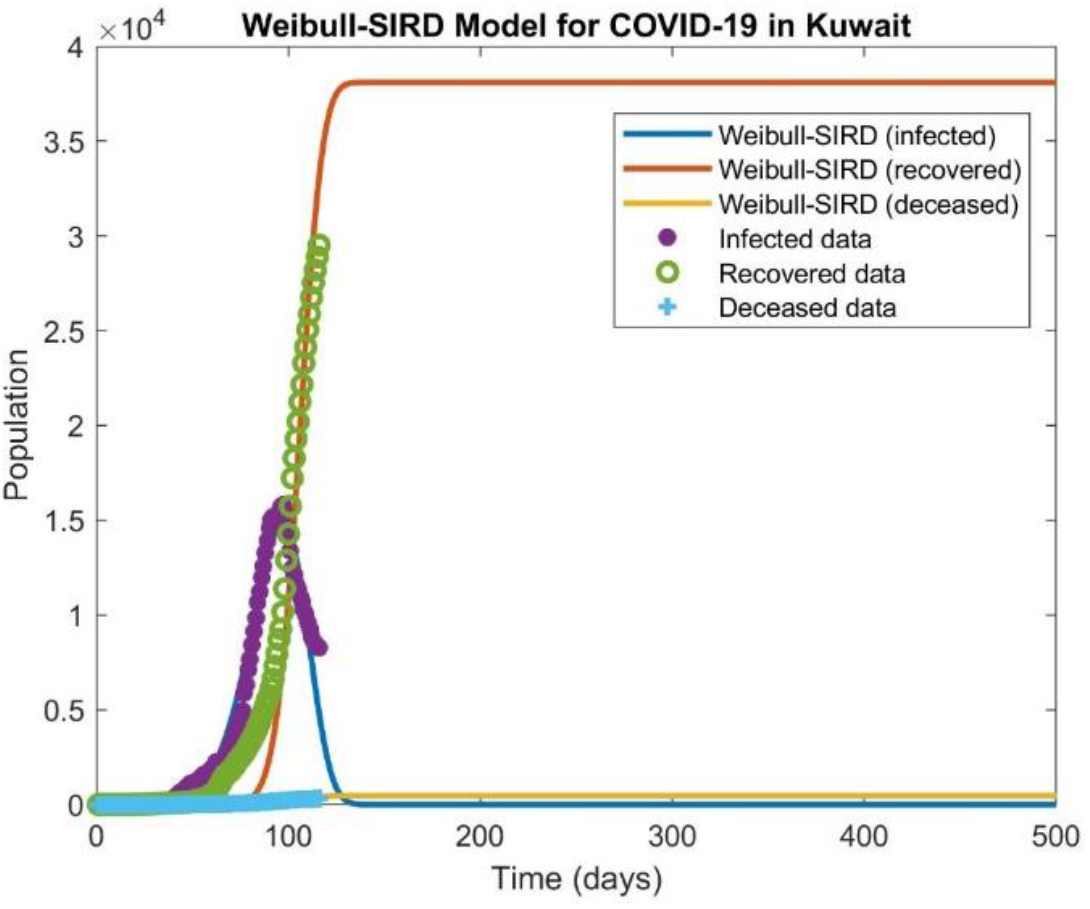
Weibull-SIRD model for fitting 4-set of COVID-19 data simultaneously in Kuwait; Top) Infected, recovered, and deceased population; Bottom) Susceptible population

### B. COVID-19 Prediction in UAE

Results of COVID-19 pandemic from 29 January 2020 to 31 May 2020 for 124 days are used to examine Weibull-SIR model. Figure 3 shows daily exposure to COVID-19 in UAE since the outbreak. Figure 4 shows Weibull-SIRD curves is simultaneously fitted with 4-set of COVID-19 data including susceptible, infected, recovered, deceased in UAE. Goodness of fit (R2) values are shown in Table 2. As shown in Fig. 4, Weibull-SIRD model predicted that on Saturday 23 May 2020 UAE had reached with the peak of COVID-19 infectious with 15,191 infected cases whilst the reported COVID-19 data was 13,769 cases on this date. The standard SIRD model has predicted 15,607 infected cases.

**Table 2:** Optimized Weibull-SIRD parameters for COVID-19 in UAE (31 May 2020).

| Total population (N) | shape factor (k) | Scale factor (c) | Growth rate ($\beta$) | Recovery rate ($\gamma_{R0}$) | Death rate ($\gamma_{D0}$) | Removing rate ($\gamma_0$) | Reproduction number ($R_0$) | $R^2(S)$ | $R^2(I)$ | $R^2(R)$ | $R^2(D)$ |
| --- | --- | --- | --- | --- | --- | --- | --- | --- | --- | --- | --- |
| 803,827 | 6.0 | 118.45 | 0.0628 | 0.0474 | 0.00085 | 0.02719 | 1.3 | 0.9927 | 0.9887 | 0.9862 | 0.7692 |

**Figure 3:**
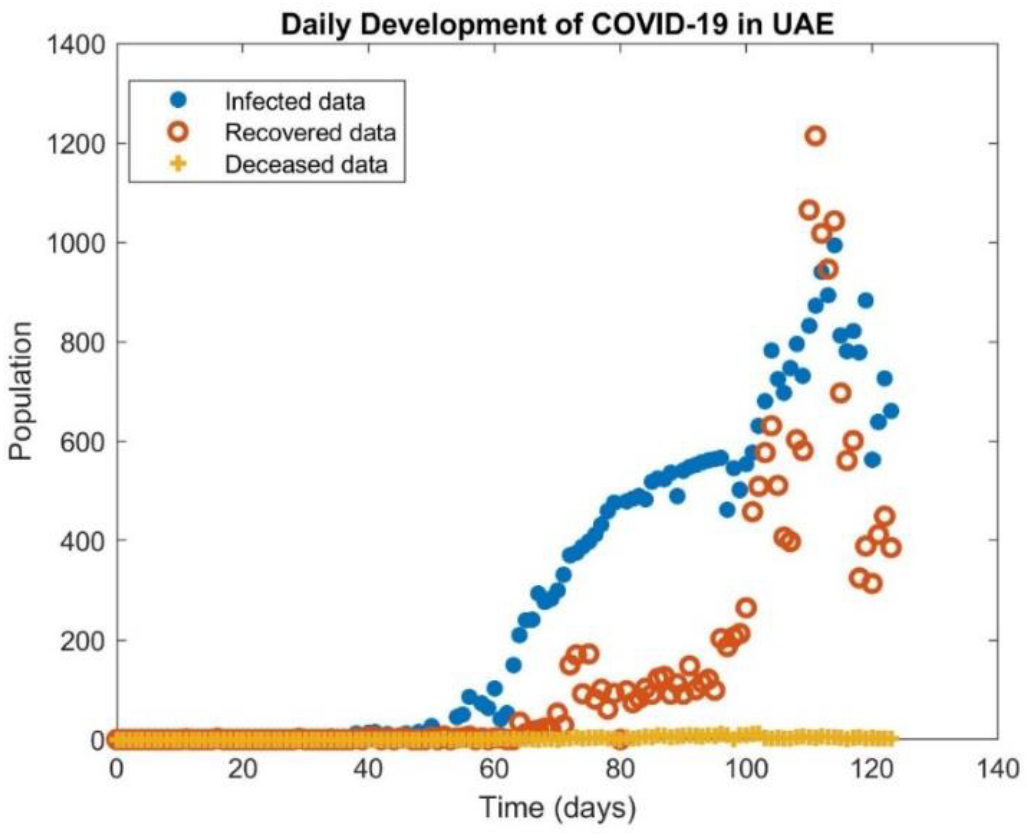
Daily exposure of COVID-19 pandemic in UAE (31 May 2020).

**Figure 4:**
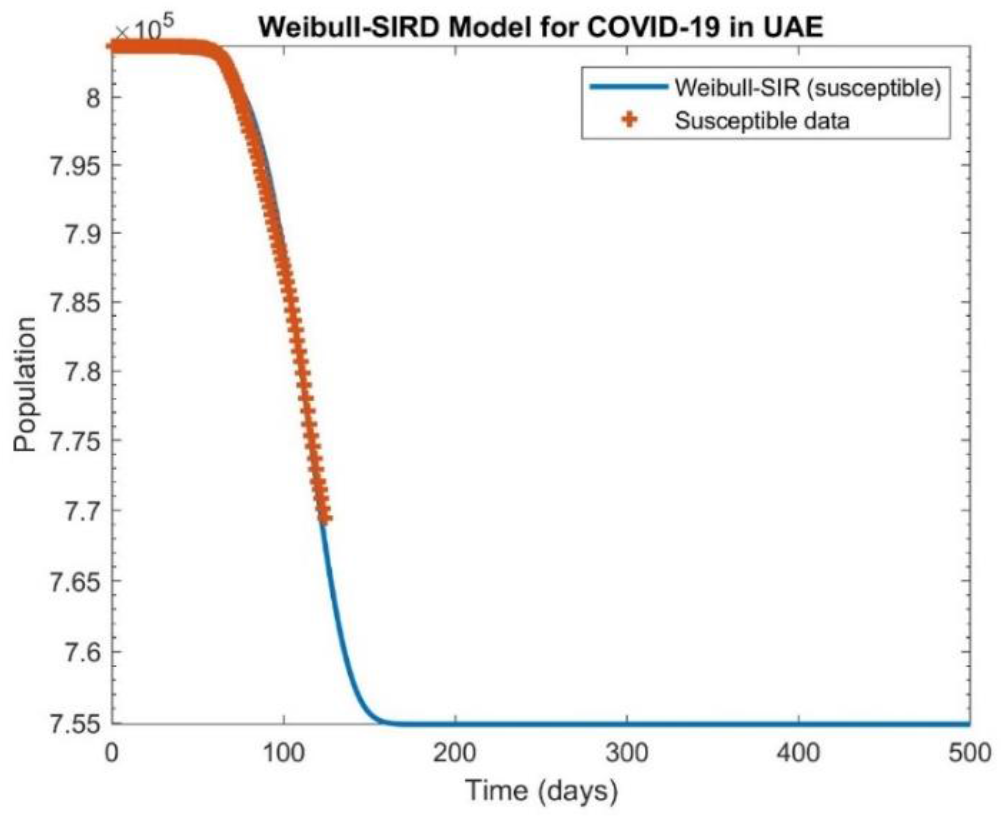
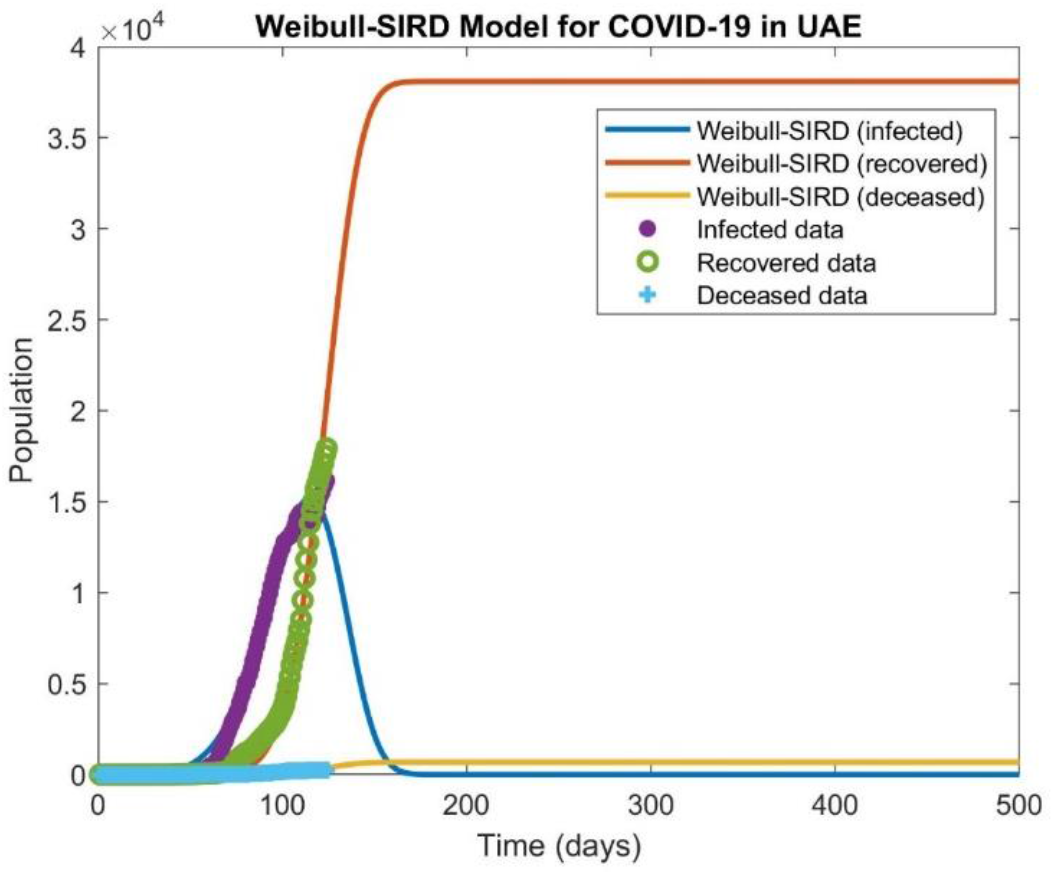
Weibull-SIRD model for fitting 4-set of COVID-19 data simultaneously in UAE; Top) Infected, recovered, and deceased population; Bottom) Susceptible population (31 May 2020).

The results of fitting equations (6) - (13) with COVID-19 data in UAE are given in Table 2. Figure 5 compares Weibull-SIRD model with the standard SIRD ODE model. Both models fitted well with recovered COVID-19 data in UAE; although, Weibull-SIRD model seems have predicted better infected cases than SIRD ODE model. SIRD Model predicted longer recovery date whilst Weibull-SIRD model have predicted shorter period. On 21 June 2020, COVID-19 data on recovered is 31,754 cases, Weibull-SIRD predicted 35,513 and SIRD model predicted 39,674. More data files and more numerical studies on COVID-19 pandemic is needed to provide solid decision but so far Weibull-SIRD model provided promising results.

**Figure 5:**
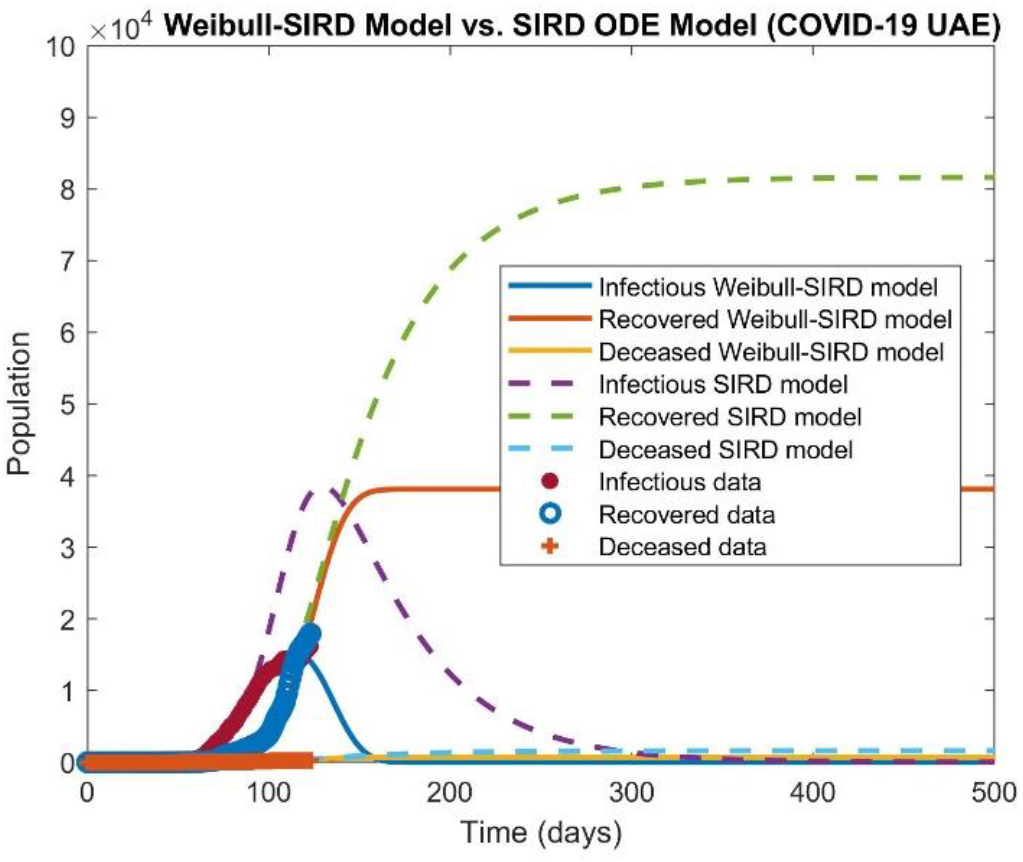
Comparison of Weibull-SIRD analytical method with SIRD ODE model for COVID-19 data in UAE (31 May 2020).

## IV. CONCLUTIONS

Better predictive methods are needed to tackle contagious diseases as early as possible. COVID-19 pandemic is a proof that we are in continuous need of better predictive methods. In this paper, a novel analytical model based on Weibull-SIRD method is introduced to study endemic/pandemic data. Weibull distribution function provides a very good probability density function to many applications in statistics and engineering including wind energy. Conclusions of this work may be drawn as follows:

- We successfully applied SIRD model to fit COVID-19 data simultaneously for infected (I), recovered (R), and deceased and suggest important peak dates and numbers of exposed populations.
- The new Weibull-SIRD model uses very simple and robust analytical formulations and fit well with COVID-19 data and is optimized for calculating model parameters.
- We studied COVID-19 data for Kuwait and UAE using both Weibull-SIRD analytical method and SIRD ODE method.
- The peak of infectious curve for COVID-19 is predicted around Sunday 31 May 2020 in Kuwait with 14,814 active infected cases.
- The peak of infectious curve for COVID-19 is predicted around Saturday 23 May 2020 in UAE with 15,191 infected cases; although, actual peak is observed on 4 June 2020 with 17,173 infected cases.

Weibull-SIRD model is a new and promising predictive method proposed here for studying endemic/pandemic outbreaks. Further works still needed to better understand all merits and features of the new Weibull-SIRD method.

## Data Availability

data is public

https://www.worldometers.info/coronavirus/

